# Non-invasive estimation of intracranial pressure by diffuse optics – a proof-of-concept study

**DOI:** 10.1101/2020.05.21.20109256

**Authors:** Jonas B. Fischer, Ameer Ghouse, Susanna Tagliabue, Federica Maruccia, Anna Rey-Perez, Marcelino Báguena, Paola Cano, Riccardo Zucca, Udo M. Weigel, Juan Sahuquillo, Maria A. Poca, Turgut Durduran

**Affiliations:** ICFO-Institut de Ciències Fotòniques, The Barcelona Institute of Science and Technology, Castelldefels (Barcelona), Spain; HemoPhotonics S.L., Castelldefels (Barcelona), Spain; Neurotraumatology and Neurosurgery Research Unit (UNINN), Vall d’Hebron Research Institute (VHIR), Universitat Autònoma de Barcelona, Barcelona, Spain; Neurotrauma Intensive Care Unit, Vall d’Hebron University Hospital, Universitat Autònoma de Barcelona, Barcelona, Spain; Department of Neurosurgery, Vall d’Hebron University Hospital, Universitat Autònoma de Barcelona, Barcelona, Spain; Synthetic Perceptive, Emotive and Cognitive Systems (SPECS), Institute for Bioengineering of Catalonia (IBEC), Barcelona Institute of Science and Technology, Barcelona, Spain; Institució Catalana de Recerca i Estudis Avançats (ICREA), Barcelona, Spain; Mediterranean Technology Park; Av. Carl Friedrich Gauss, 3; 08860 Castelldefels (Barcelona); Spain; Paseo Vall d’Hebron, 119-129; 08035 Barcelona; Spain; Baldiri Reixac, 10-12; 08028 Barcelona, Spain

**Keywords:** non-invasive, intracranial pressure, diffuse optics, near-infrared spectroscopy, neural networks

## Abstract

Intracranial pressure (ICP) is an important parameter to monitor in several neuropathologies. However, because current clinically accepted methods are invasive, its monitoring is limited to patients in critical conditions. On the other side, there are other less critical conditions where ICP monitoring could still be useful, thus there is a need to develop non-invasive methods. We propose a new method to estimate ICP based on the analysis of the non-invasive measurement of pulsatile, microvascular cerebral blood flow with diffuse correlation spectroscopy. This is achieved by training a recurrent neural network using only the cerebral blood flow as the input. The method is validated using a 50% split sample method using the data from a proof-of-concept study. The study involved a population of infants (n=6) with external hydrocephalus (initially diagnosed as benign enlargement of subarachnoid spaces) as well as a population of adults (n=6) suffering from traumatic brain injury. The algorithm was applied to each cohort individually to obtain a model and an ICP estimate. In both diverse cohorts, the non-invasive estimation of ICP was achieved with an accuracy less than <4 mmHg and a negligible small bias. Furthermore, we have achieved a good correlation (Pearson’s correlation coefficient >0.9) and good concordance (Lin’s concordance correlation coefficient >0.9) in comparison to standard clinical, invasive ICP monitoring. This preliminary work paves the way for further investigations of this tool for the non-invasive, bed-side assessment of ICP.

## Introduction

Intracranial pressure (ICP) is a biomarker that is routinely monitored in many neurocritical care patients and in certain types of disorders of cerebrospinal fluid (CSF) dynamics. Typically, ICP is measured invasively by implanting an intracranial sensor through a burr hole or a twist drill in the skull. However, this procedure is associated with a risk of infection and/ or hemorrhagic complications.^1^ Since the procedure is invasive, ICP is only monitored in patients suffering from critical pathologies such as traumatic brain injury (TBI), subarachnoid hemorrhage and difficult cases of hydrocephalus where the benefit of continuous ICP monitoring outweighs the risks. For example, continuous, invasive ICP monitoring is recommended in patients with severe TBI since it has been established that, as a secondary injury, raised ICP is a cause of neurological deterioration and is related to an unfavorable neurological outcome.^2^ The monitoring of ICP allows the clinicians to manage elevated ICP via a set of interventions. In other conditions such as difficult cases of hydrocephalus, ICP monitoring helps clinicians to decide whether a CSF shunt is required or not.

As another use case, ICP monitoring may also be relevant for the management or diagnosis of other neurological conditions in different populations, especially in pediatric populations,^3^ in patients affected by certain pathologies such as acute liver failure^4^ or in patients suffering from moderate TBI, large hemispheric ischemic stroke or aneurysmal subarachnoid hemorrhage. However, routine ICP monitoring is not recommended in these cases due to its aforementioned risks. To overcome this limitation, over the past 40 years, several non-invasive techniques of ICP monitoring have been proposed but no method has been accurate enough to be widely implemented in clinical practice.^1,5,6^

The spontaneous fluctuations of the pulsatile cardiac component of the cerebral blood flow (CBF) is often considered as a non-invasive surrogate of ICP.^1,5,6^ To that end, transcranial Doppler ultrasound (TCD) has been widely utilized as a tool to try to estimate these changes based on the measurement of the blood flow velocity in a major artery. A detailed overview of those techniques can be found in Cardim et al.^7^ where the use of simple parameters such as the pulsatility index (PI) — the difference between the systolic and diastolic blood flow velocities normalized by the mean blood flow velocity — have been reported alongside more advanced models and methods including machine learning models, both in the supervised and unsupervised sense, to predict the cerebral perfusion pressure (CPP). We note that most of these models require a reliable, beat-to-beat measurement of arterial blood pressure (ABP) as an input parameter.

Unfortunately, despite major advances in head-gear and robotic control of the transducers, TCD is not yet a practical technique due to its operator dependence, motion sensitivity and the lack of a usable bone window insonation in a significant number of the patients.^7,8^ The need for non-invasive, practical ICP monitoring is still unmet and it has motivated researchers to explore the potential of diffuse optical techniques such as near-infrared spectroscopy (NIRS)^9^ as other alternative surrogate measures.

NIRS was first used in the neurocritical care as a monitor for detecting the desaturation of the cerebral blood oxygenation with a better time-resolution compared to the widely used venous saturation in the jugular bulb (SjO_2_).^10^ Since cerebral extraction of oxygen drops when CBF is reduced and raised ICP reduces CPP and CBF, brain oxygen desaturation is an indirect but imprecise estimator of high ICP. It has also been shown that changes in the NIRS signal indicating hemoglobin desaturation are related to changes in the ICP.^11,12^ However, the NIRS approach is limited because it is only an indirect index of episodes of raised ICP that influence brain oxygen desaturation and it is also heavily influenced by other variables apart from ICP (mean arterial blood pressure MABP, arterial oxygen saturation SaO_2_, hemoglobin content, and others). Typical NIRS measurements do not allow for absolute measures of ICP and, therefore, its clinical adoption requires building up new evidence and guidelines since the current guidelines for conditions such as the severe TBI management algorithms are based on absolute ICP thresholds.^13^

There is an emerging complementary approach to traditional NIRS, diffuse correlation spectroscopy (DCS), which is also a near-infrared, diffuse optical technique that instead measures a blood flow index (BFI) in a non-invasive and continuous manner to directly retrieve the microvascular CBF by quantifying the laser speckle statistics.^14^ Recent progress in DCS technology allows for fast measurements (~10-100 Hz) capable of resolving the pulsatile behavior of blood flow due to the cardiac cycle.^15,16^ It has been suggested, in a similar manner to the TCD methods, that one can now utilize the beat-to-beat variations in the CBF signal with additional beat-to-beat information of the ABP to estimate the critical closing pressure (CrCP) as a surrogate for the ICP.^17,18^ Preliminary results have shown a good correlation of DCS measures with ICP values in infants suffering from hydrocephalus.^19^ The main disadvantage of this approach is that continuous measurements of the CrCP require continuous and reliable ABP measurements which are often not readily available.

In our work, we have been motivated by the fact it is well known from previous work with TCD, that the CBF waveform is significantly altered due to the external influences of the ICP on the vasculature with a complex and non-linear relationship.^20^ In fact, previous studies have shown that ICP and intracranial compliance can be estimated by TCD based measures^21^ or by using the waveform morphology of the pulsatile ICP signal using machine learning methods.^22,23^ Therefore, we have hypothesized that a nonlinear, autoregressive, machine learning model can identify a system that maps the pulsatile CBF onto the ICP. A recurrent neural network (RNN), which is a class of neural networks is a suitable method for this type of time dependent data, was selected for our purpose.

Here, we present data of a proof-of-concept study in which we have employed an RNN based method that maps the pulsatile, microvascular CBF that was measured non-invasively by DCS to the invasive ICP measurements conducted by conventional intracranial extradural or intraparenchymal ICP sensors. We demonstrate the capabilities of the model in a population of infants with external hydrocephalus and in another population of adults suffering from severe TBI.

## Methods

### Clinical populations

#### Pediatric population

This part of the study was conducted at the Neurosurgical Pediatric Unit of the Vall d’Hebron University Hospital (Barcelona, Spain). The study protocol was approved by the local ethical committee (PR(AMI)459/2017). Infants aged between 0 and 42 months, initially diagnosed with the “benign” enlargement of subarachnoid spaces (BESS) were recruited for the study. The parents of the infants gave their informed written consent.

Patients diagnosed with BESS presented a combination of macrocephaly or an abnormal acceleration in the growth of the head circumference along with associated clinical symptoms such as delays in psychomotor development and irritability. All patients had a moderate dilation of the ventricular system. ICP was continuously monitored using an extradural device to confirm the diagnosis of external hydrocephalus and decide if the child requires a CSF shunt. A DCS probe was applied to the frontal lobe on the same hemisphere where the ICP probe was placed for a minimum of thirty minutes after the infant fell asleep.

#### Adult population

The second part of the study was conducted at the Neurotrauma Intensive Care Unit of the Vall d’Hebron University Hospital (Barcelona, Spain). The study protocol was approved by the local ethical committee (ACU-AT-203/2012(3531)). The written informed consent was obtained from the next-of-kin of the patients. Patients admitted to the neurotrauma care unit older than 18 years with a moderate or severe TBI (admission Glasgow Coma Score ≤ 13) requiring mechanical ventilation and ICP monitoring were eligible for this study. Pulsatile CBF was measured for at least thirty minutes on one frontal lobe, usually on the same hemisphere where the cranial sensors were implanted (parenchymal ICP, brain tissue oxygen pressure (PtiO_2_) and cerebral microdialysis). The presence of an open wound, extensive sutures, craniectomy and other issues that hinder reliable optical measurements or the probe placement was the reason for the utilization of the contralateral hemisphere for the measurements. All patients were sedated at the time of the measurement. We note that these patients were enrolled for a larger study related to non-invasive transcranial, bed-side monitoring of hemodynamics with optics. Therefore, some of the data recorded was taken during patient manipulations, such as changes in head-of-bed position, or during a protocol of hyperventilation, as indicated in the results.

Both studies were carried out following the principles of the Declaration of Helsinki.

### Optical method and instrumentation

A DCS system^14^ was developed for this study consisting of a long coherence laser at 785 nm (iBeam smart WS, Toptica Photonics AG, Germany), eight single-photon avalanche diodes (2x SPCM-AQ4C, Excelitas Technologies, Canada) and a custom hardware correlator (CM 8, HemoPhotonics S.L., Spain). The custom hardware correlator allows for fast measurements up to 100 Hz but the acquisition rate was set to ~40 Hz to ensure a sufficient signal quality while still being able to sufficiently resolve the details of the pulsatile blood flow.

A custom made optical probe with a source-detector separation of 2.5 cm was developed. The probe consisted of a foam piece as the main body and incorporated a 90° bent multi-mode fiber (Fiberoptic Systems Inc., USA) for the source as well as a 90° bent fiber bundle (Fiberoptic Systems Inc., USA) with seven single-mode fibers for the detection of the light.

The time-course of the CBF was obtained by applying the modified Beer-Lambert law for blood flow.^24^ The signal-to-noise ratio was increased by averaging the signal of the seven channels.^15^ The first five minutes of each measurement were used as a baseline for the analysis. In order to increase the sensitivity to the brain, we have considered only early delay times of the normalized electrical field correlation function *g*_1_ for the calculation of the BFI following *g*_1_ > 0.63 for an average correlation curve of the baseline.^25^ As optical properties, we have assumed a reduced scattering coefficient of 10 cm^−1^ and an absorption coefficient of 0.1 cm^−1^ for all the analysis. The calculated BFI time course was used as the input to the neural network.

### Intracranial pressure sensors and signal synchronization

In the pediatric patients with suspected external hydrocephalus, the ICP was measured with an epidural sensor (NEURODUR-P®, Raumedic AG, Germany). In the TBI population, the ICP was measured using an intra-parenchymal sensor (Neurovent-PTO®, Raumedic AG, Germany or Camino®, Integra LifeSciences, USA). Both were part of the routine clinical care of the patients.

In both cases, the optical signals and the ICP measurements were synchronized using the LabChart software v7.0.3 (ADInstruments, New Zealand) and the data acquisition hardware PowerLab (ADInstruments, New Zealand). The ICP signal was directly fed from the ICP monitor to PowerLab and the correlator sent a 10 Hz digital signal to PowerLab as a timing basis.

### Learning architecture and algorithm

The algorithm and the neural network were implemented in Python 3.5.2 and PyTorch 1.0.0. The model was trained on a GeForce GTX-1080 GPU (Nvidia Corporation, USA) with eight gigabytes of memory.

As previously noted, we have used an RNN to model a system generating an ICP estimate using the time series of the pulsatile CBF as an input. The results from a gated recurrent unit (GRU)^26^ architecture are reported due to reduced memory and computational restraints with a similar performance as other architectures such as long short term memory (LSTM) networks.^27,28^

Due to the limited number of the patients that were planned, we have used a 50% holdout validation method, otherwise known as random sample splitting validation. Here, the 50% of the data was randomly assigned for training, and, the other 50% was used for testing. The algorithm was implemented on a windowed basis, meaning that windows of data – as detailed below – from a subject’s time course of pulsatile CBF were used as the input to the neural network and an ICP estimate was predicted for each window.

The general flowchart for training and testing our model is shown in Figure 1. The data was first pre-processed by low pass filtering the raw data (sampling rate ~ 40 Hz). The filter had a cut off frequency of 7 Hz in order to capture at least three harmonics of the heart rate while retaining low frequency data such as the fluctuations induced by the respiratory rate. The same filtering method was applied to the ICP data. Then, the entire filtered time course of the CBF with *N_M_* measurements was windowed in *N_W_* sections of several cardiac cycles with a length *L_W_* of 600 measurements or roughly fifteen seconds given the acquisition rate of ~40 Hz for one measurement. A moving window was performed with a stride of *L_stride_* = 25 measurements. This was done to maximize the independence between the initial time point of each window and to attempt to retain a data-set that is relatively independent of the initial states. This leads to a window sampling rate of ~1.6 Hz. The center of each time window was labeled with a new time-base where the number of windows *N_W_* is given by *N_W_* = *N_M_*/*L_stride_*. Finally, the data range for each window was standardized such that the input data in the RNN would have a zero mean and a standard deviation of one. This normalization ensures that the network does not bias its learning based on the shifts of the mean level of the data. The goal is to have the network learn the features of the waveform of the pulsatile CBF itself without being heavily influenced by potential variations of the mean CBF. The ICP data was windowed using the same method adopted to the sampling rate of the ICP (200 Hz or 400 Hz depending on the sensor) and the average value of each ICP window was calculated serving as the labels, i.e. the ground-truth value for the training algorithm. The random sampling method for the training algorithm is described in detail in the Supplementary Appendix S1.

**Figure 1:**
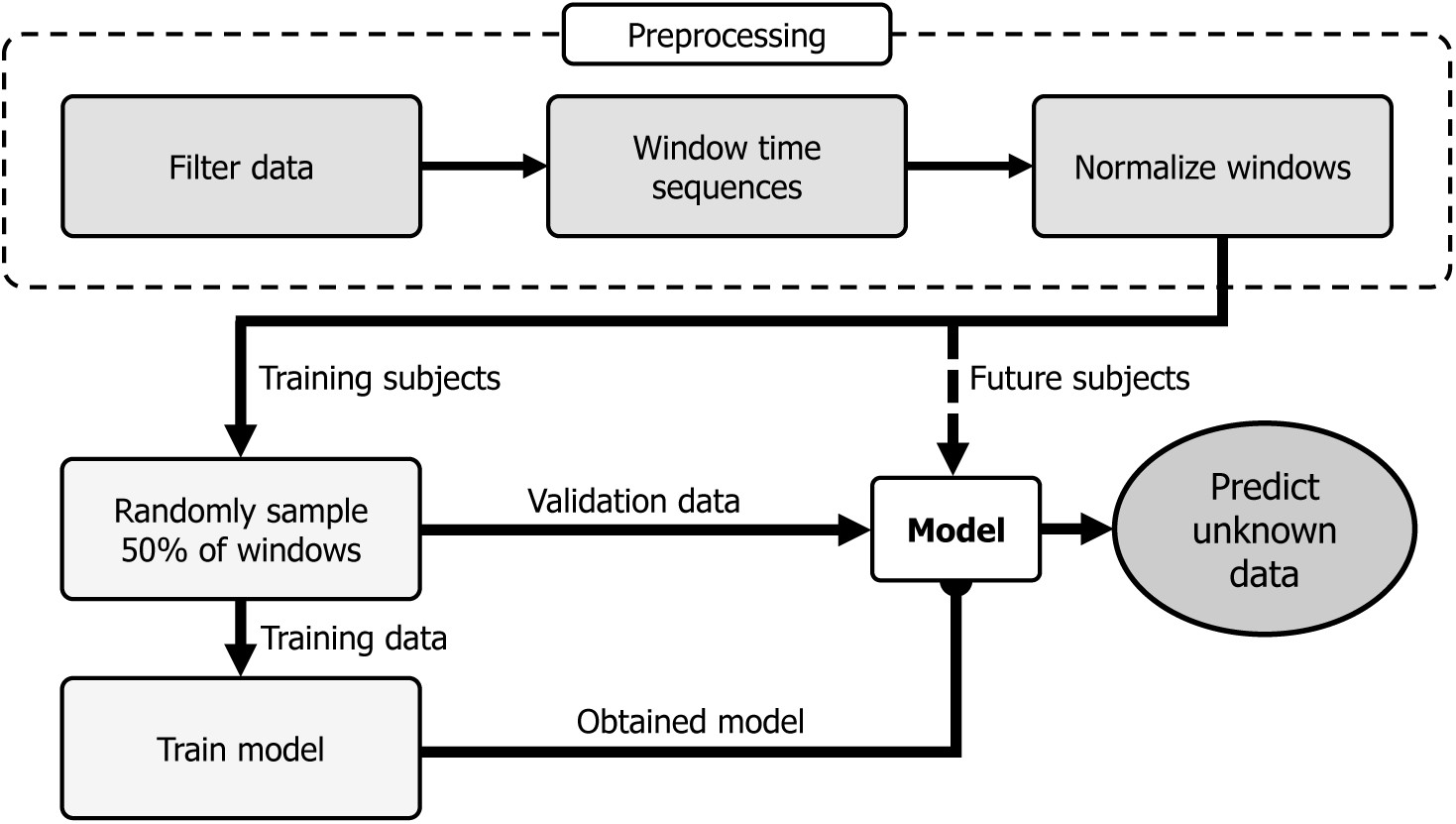
Algorithm flowchart. After pre-processing, the model is trained with up to 50% of the data as the training set to simulate a “weak learner”. The validation data and additional subjects can be evaluated by applying the obtained model. The normalized windows of the pulsatile CBF are the input to the model. See text for further details.

The general architecture of the RNN model and how the windows are fed into the network is shown in Figure 2. All hyperparameters of the network were empirically determined. The pre-processed windows with a length of *L_W_* shown in blue are subsequently passed into the algorithm in subwindows (*x*(*t*′)) with a length of 20 measurements shown in red. The input layer is then passed into a multi-layered RNN. In our particular case, we have used a GRU RNN consisting of two layers with sixty-four hidden states in each layer. For each time step, *t*′, the hidden states *h_x_*(*t*′) are updated. Finally, at the last step of a window, a linear layer is used as a regression layer to output a prediction for a window *i*. The predicted value *y_i_* can then be compared to the ICP label, which is the average of said window 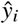. For the training, the mean squared error (MSE) was used to determine the error between the prediction *y_i_* and the result found by the given ICP labeled data 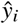, as 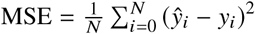, where *N* is total number of the windows used for all subjects.

**Figure 2:**
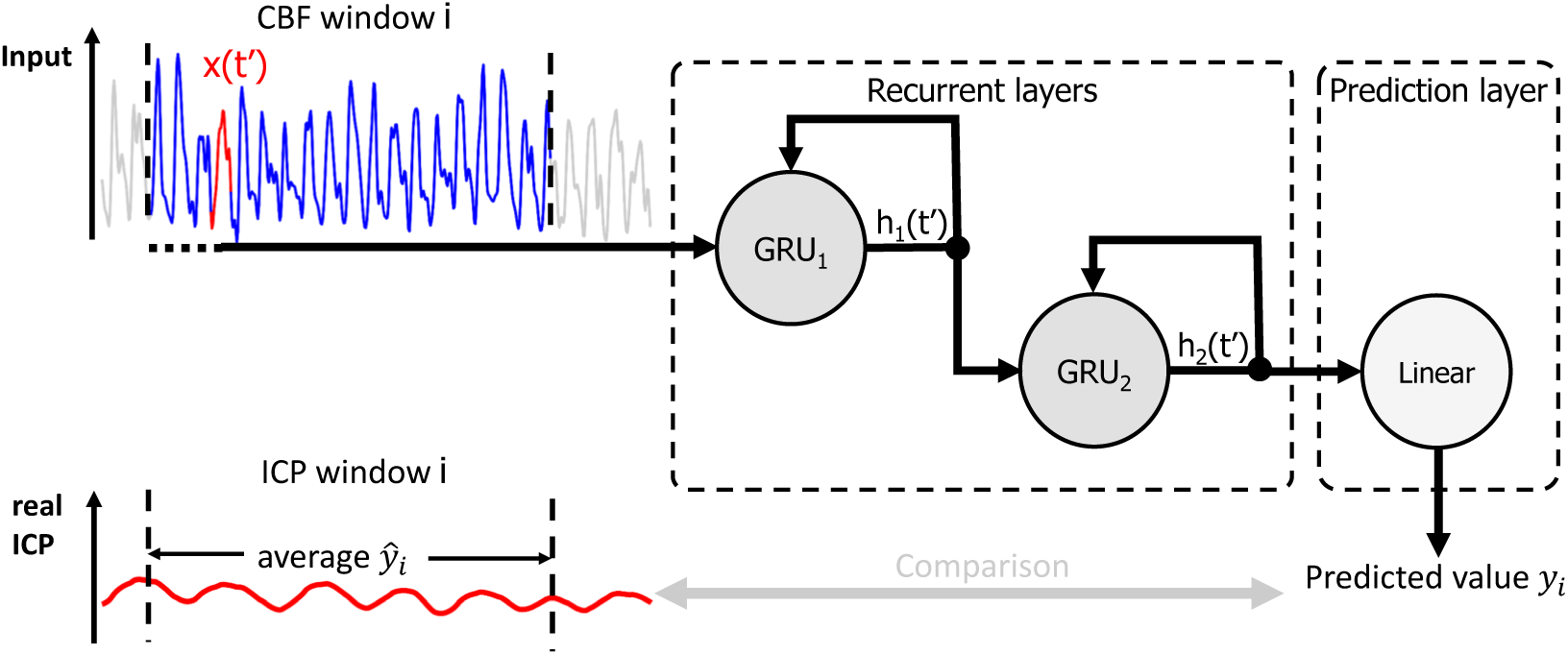
Schematic structure of the RNN. A window (blue) of the pulsatile CBF is fed sequentially with subwindows (red) into the input layer. The RNN consists of a multi-layer structure with two GRU cells followed by a prediction layer with a linear cell to predict one ICP index for the said CBF window. This index can be then compared to the average of the same window of the ICP time series, which can also act as a label (a ground-truth value for a certain set of feature) when the model is trained. The definitions for *h_x_(t′*), 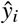 and *y_i_* can be found in the text.

Back-propagation, which is the algorithm to calculate the gradient of the error function, was used at the last time step of a window to calculate the gradient of the error with respect to the weights of the network. This allowed for stochastic gradient descent, particularly the Adam algorithm, to optimize the weights of the neural network.^29^ Here, we have used a learning rate of 10^−3^, which is a parameter that controls how fast the model learns by setting a fixed amount of change for each step in the process. One-hundred epochs were used to train the model, where one epoch represents the full presentation of the training set to the network to update the weights. The learned weights of the network were stored for the analysis of the validation data set. The predicted ICP time series was binned in 30 second bins (~ 45 windows).

### Statistical analysis

The analysis of the data was conducted with MATLAB (MathWorks, USA). For the statistical analysis we report results only from data that was included in the validation set. Each data point represents an average value derived from the aforementioned 30 second bin for both invasive and non-invasive data. In cases where the training and the validation data appear in the same 30 second time bin (due to the random sampling), the majority (i.e. > 50%) was identified and used for assigning the bin to the particular group.

We have checked for the correlation between the non-invasive estimation of an ICP with the real measured ICP. The slope and the intercept are reported with their 95% confidence interval (95% CI). Furthermore, we present the Pearson’s (***R***) correlation alongside Lin’s concordance correlation coefficient (*ρ_c_*) which is a measure how well a new method agrees with a “gold standard” method.^30^

A Bland-Altman analysis was used to to check for the agreement of both methods by looking at the differences between the DCS method and the gold standard.^31^ In particular, we have calculated the Bland-Altman plot for the differences as a percentage ([non-invasive ICP – invasive ICP] / mean ICP x 100%) in order to detect potential systematic bias at low or high ICP values.

For a more complete picture of the data, we provide supplementary materials with further subject specific information such as the distribution of the ICP for each subject as well as all the time traces showing the ICP measured with the invasive sensor compared to the non-invasively estimated ICP.

## Results

### Benign enlargement of subarachnoid spaces cohort

Six infants with external hydrocephalus (initially diagnosed with BESS) were included in the study. The median age of the population was 26.5 months (min: 7, max: 55 months)*^a^*. Time series of CBF was obtained alongside ICP measured in the epidural intracranial space of the left hemisphere. Table 1 summarizes the demographic data and the methodology used in each patient. Furthermore, the ICP results are further detailed in the Supplementary Appendix S2 in Figure S1 where histograms of the measured ICP values are shown for each subject in order to assess the distribution of values available for training the algorithm.

**Table 1:** Overview of the BESS population including information of the measurement.

| ID | Gender | Age | Location | Monitored time |
| --- | --- | --- | --- | --- |
|  |  | [months] | DCS | [min] |
| B1 | M | 23 | left | 30 |
| B2 | M | 30 | left | 32 |
| B3 | M | 55 | left | 60 |
| B4 | M | 11 | left | 32 |
| B5 | F | 7 | left | 30 |
| B6 | F | 33 | left | 31 |

In Figure 3a (also Supplementary Appendix S2, Figure S2a), the predictions from the validation set of time windows are plotted against the readings of the invasively measured epidural ICP sensor (see Supplementary Appendix S2, Figures S3 to S8 for individual time traces). The linear regression slope was 0.85 (95% CI: 0.82, 0.89) and the intercept was 2.8 mmHg (95% CI: 2.1 mmHg, 3.5 mmHg). The Pearson correlation coefficient was *R* = 0.95 and the Lin’s concordance correlation was *ρ_c_* = 0.95. In order to analyze the agreement between the two methods, a Bland-Altman analysis was performed, comparing the differences as a percentage in Figure 3b (also Supplementary Appendix S2, Figure S2b). The mean difference, or bias, between the prediction and gold standard was +0.82 %, corresponding to a +0.15 mmHg difference. The standard deviation was 8.0%, yielding an accuracy (95% CI) of ±3.0 mmHg.

**Figure 3:**
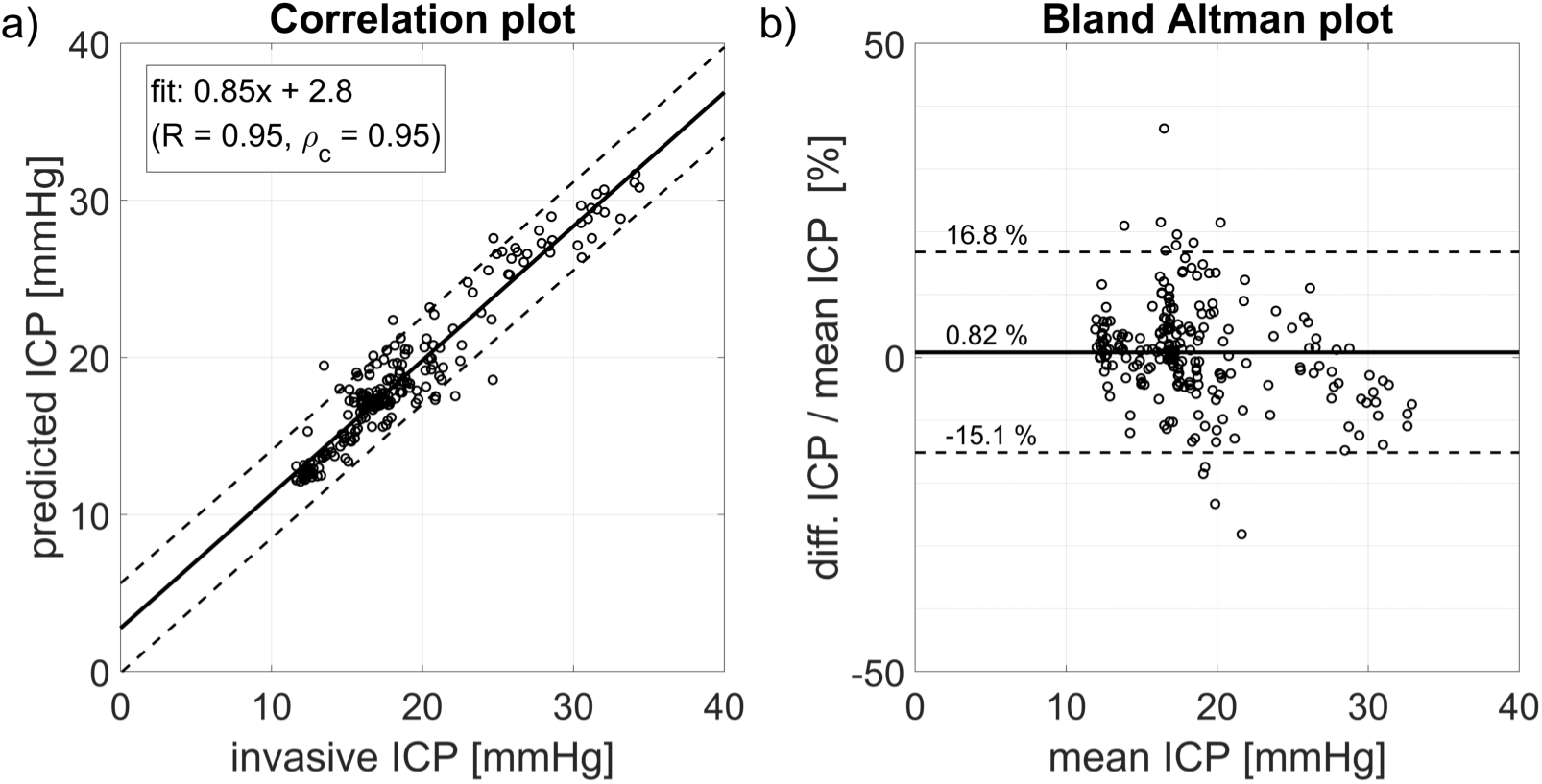
BESS subjects: Panel a) Correlation analysis. The solid line represents the best linear fit and the dashed lines the 95% confidence interval of the fit. Panel b) Bland-Altman plot. The solid line represents the mean (= bias) and the dashed lines the mean ± 1.96 standard deviations. (mean ICP = mean of predicted ICP and invasive ICP, diff. ICP = predicted ICP - invasive ICP). See text for more details.

### Traumatic brain injury cohort

Six patients with a moderate or severe TBI that required ICP monitoring were recruited. The median age of this population was 35.5 years (min: 21, max: 39 years). Table 2 reports a summary of the subjects and the clinical settings. We note that Subjects 3 and 4 have repeated measurements at different days. The histograms of the ICP measurements for each subject can be found in Supplementary Appendix S3 in Figure S9 as before.

**Table 2:** Overview of the TBI population including information of the measurement. (R denotes Raumedic, C denotes Camino and parench. stands for intraparenchymal.)

| ID | Gender | Age<br>[years] | Type of<br>ICP sensor | Location<br>ICP | Location<br>DCS | Monitored time<br>[min] |
| --- | --- | --- | --- | --- | --- | --- |
| T1 | M | 39 | parench. (R) | left | right | 42 |
| T2 | M | 34 | parench. (C) | right | right | 30 |
| T3 (1) | M | 39 | parench. (R) | right | right | 87 |
| T3 (2) |  |  |  |  |  | 170 |
| T4 (1) | M | 37 | parench. (R) | right | left | 180 |
| T4 (2) |  |  |  |  |  | 180 |
| T5 | M | 23 | parench. (R) | right | right | 152 |
| T6 | F | 21 | parench. (R) | left | left | 30 |

The linear regression of the predicted versus measured ICP values for the TBI patients is shown in Figure 4a (also Supplementary Appendix S3, Figure S10a). Furthermore, for clarity, Figures S11 to S18 in the Supplementary Appendix S3 show individual time traces. The slope of the linear regression was 0.90 (95% CI: 0.88, 0.92) with an intercept of 1.2 mmHg (95% CI: 0.94 mmHg, 1.4 mmHg). The Pearson’s correlation coefficient was *R* = 0.96 and the concordance correlation was *ρ_c_* = 0.96. The differences as a percentage between the two methods were analyzed by the Bland-Altman plot which is shown in Figure 4b (also Supplementary Appendix S3, Figure S10b). The bias between the two methods is +0.69 % which corresponds to +0.087 mmHg. The standard deviation was 13.4% leading to an accuracy (95% CI) of ±3.4 mmHg.

**Figure 4:**
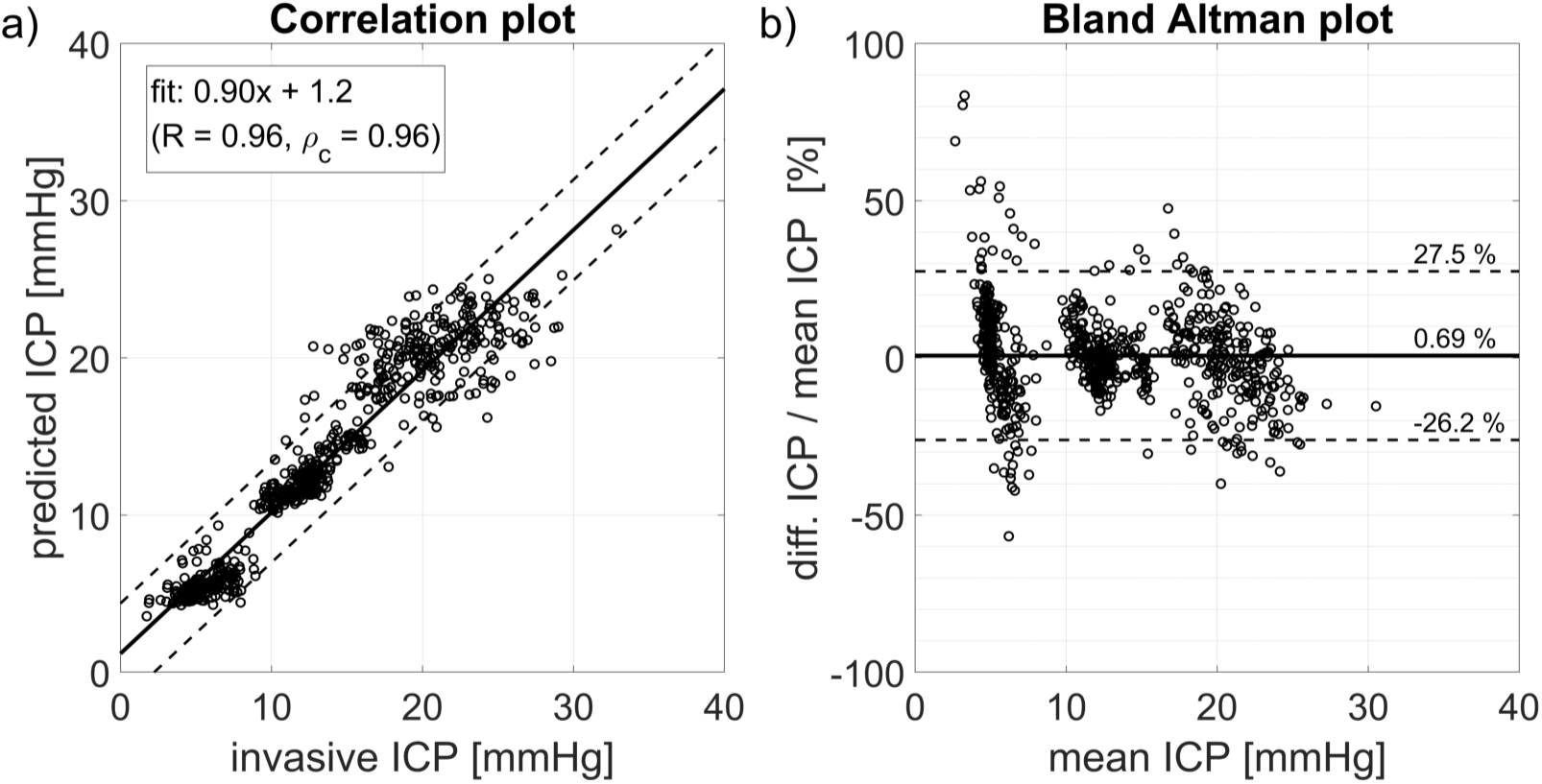
TBI subjects: Panel a) Correlation analysis. The solid line represents the best linear fit and the dashed lines the 95% confidence interval of the fit. Panel b) Bland-Altman plot. The solid line represents the mean (= bias) and the dashed lines the mean ± 1.96 standard deviations. (mean ICP = mean of predicted ICP and invasive ICP, diff. ICP = predicted ICP - invasive ICP). See text for more details.

## Discussion

We have described and introduced a new method to non-invasively predict ICP based on the optical measurement of pulsatile CBF with DCS together with an RNN model. Overall, we were able to map the pulsatile CBF to the ICP readings of the invasive sensors with a machine learning algorithm both in infants and in adults (See Figures 3 and 4).

We highlight here that the algorithm gave promising results for the two different models in infants with a mean age of 26.5 months as well as in adults with a mean age of 32 years. This is remarkable, since the two populations have a different head anatomy due to their age difference. In particular, the depth of the brain from the scalp surface is different which will alter the partial volume effect^32^ and the origin of the optical signal. In other words, the fraction of the DCS signal that is coming from the brain can be significantly different.^25^ The same optical probe with a source-detector separation of 2.5 cm was used in both populations meaning that we were more sensitive to the brain in the infants due to the thinner skull.

Nonetheless, the algorithm gave similar results for the adult population. We speculate that the fact that the algorithm performed well for adults and infants with different head geometries is because the predicted ICP is related to the pulsatile component of the CBF. In order to prove that the information about the ICP is mainly contained in the pulsatile dynamics of the CBF, we have trained the algorithm on a low-pass filtered data set (0.4 Hz) of the BESS cohort yielding a poorer correlation (*R* < 0.5) and a poorer concordance (*ρ_c_ <* 0.35) which are well below being acceptable.

Taken together, these findings can be seen as an indication that the pulsatility has an enhanced sensitivity to deeper layers, meaning in particular to the cortex compared to the extracerebral layers. This speculation is further strengthened by the findings of a recent study where the authors have shown that the pulsatility of the DCS measured CBF is not affected by the probe pressure.^33^ This finding also implies enhanced sensitivity to the deeper layers for the pulsatile component compared to traditional, low frequency measurements where probe pressure was shown to modulate the DCS signal.^34,35^

Head anatomy is not the only factor that could have led to differing results in populations since different ICP sensors introduce different systematic errors in estimating the true ICP. For example, the extradural ICP sensors (used in the BESS group) are known to overestimate the real ICP compared to parenchymal or ventricular ICP sensors in adults.^36,37^ Therefore, we have chosen to train different models per population set.

Different populations have different needs as well. Invasive ICP monitoring is not common in pediatric BESS patients since is often considered a self-limiting condition that does not require treatment. Our clinical partners were motivated to carry out ICP monitoring in BESS patients due to recent literature (and yet unpublished data) demonstrating that BESS is associated in some cases to a persistent delay in motor and cognitive skills.^38^ The hypothesis here is that in some of these patients persistent alterations may exist in the CSF dynamics which could be demonstrated by continuous ICP monitoring. If our method for non-invasive optical monitoring for estimation of ICP is validated, it may shed light into this line of unknowns leading to changes in clinical practice.

In TBI patients, the most relevant information is the mean ICP value in order to detect intracranial hypertension. Typically, non-invasive ICP methods are assessed by performing a receiver operating characteristic (ROC) analysis for a threshold of 20 mmHg, to get a measure of their ability to detect raised ICP. If we assume that our model will generalize to out-of-sample subjects in a similar manner, an analysis of the ROC curve for the detection of ICP values above 20 mmHg for the TBI population could be evaluated to show its usefulness in the neuro-critical care setting. We have carried out an exercise despite the limitations (see below) of our study to evaluate its performance and an area under the curve (AUC) of 0.96 with an optimal threshold of 17.5 mmHg for the DCS estimated ICP showed a sensitivity of 96% and a specificity of 88%. Furthermore, we have calculated performance measures for the said threshold which are summarized in Table 3. This illustrative exercise of taking the small data set and calculating performance measures motivates us to obtain a larger training data-set and evaluate the potential of the method for online, non-invasive detection of elevated ICP.

**Table 3:** Performance measures to detect raised ICP (20 mmH used as a threshold) based on the estimated ICP by the RNN. Only validation data was taken into account. (PPV = positive predictive value; NPV = negative predictive value)

| Performance measure | >20 mmHg |
| --- | --- |
| Sensitivity | 75% |
| Specificity | 95% |
| PPV | 85% |
| NPV | 91% |
| Accuracy | 89% |

We note that these results are in line with other studies. Kashif et al.^39^ obtained an AUC of 0.83 with a sensitivity of 83% and a specificity of 70% for detection of elevated ICP based on a threshold of 20 mmHg and their model based estimation of ICP. Kim et al.^21^ reported an AUC of 0.92 using a semi-supervised method in a heterogeneous cohort of TBI, aneurysmal subarachnoid hemorrhage and normal pressure hydrocephalus patients.

Moreover, a non-invasive ICP monitor ideally should not only detect elevated ICP but also be sensitive to small changes even though they might not be clinically relevant. For example, if a controlled alteration of the patient status causes an expected change, the clinical care staff may use this as a means to ensure the correct functioning of the probe. In our case, a head-of-bed (HOB) position change (HOB angle of 25° → HOB angle of 10° for ~15 min → HOB angle of 25°) was captured in one subject showing a continuous measurement of the expected ICP changes as shown in Supplementary Appendix S3, Figure S16. This is of importance for a device which provides critical information such as the ICP, such that the clinicians can trust its output. On the contrary, one may develop a device which only detects periods of elevated ICP instead of providing a continuous ICP information. Further studies will allow us to evaluate the true potential.

Another relevant point for TBI management is the temporal resolution. According to the BrainIT consortium, one mean ICP value per minute is sufficient for critical care patients suffering from a TBI^40^ which compares well to our thirty second time-bins. On the other hand, in hydrocephalus patients, qualitative information such as the presence of ICP waves is of importance. ICP B-waves occur with frequencies of up to 3 cycles per minute,^41^ which would require a higher temporal resolution for the prediction of an ICP index. A larger training data set should reduce the the prediction error on unseen data, thus allowing us to tune the time resolution parameters of the algorithm in order to get a better temporal resolution with potential equivalent or better performance. In particular, the binning parameter can be adjusted to obtain a temporal resolution of one cycle per second so that ICP waves can potentially be detected. However, slow wave oscillations might also occur in TBI patients. In one patient of the TBI cohort we observed visibly recognizable slow waves which were caught by the DCS estimated ICP (see Supplementary Appendix S3, Figure S14).

One of the limitations of our study was that the sample size of recruited subjects for both populations was small and the measurement periods were relatively short implying that a lot of our data was derived from periods of relatively stable ICP measurements. One can observe this in individual time-traces and histograms shown in the Supplementary Appendix S2 and Supplementary Appendix S3. This has hindered us from carrying out an analysis for each individual such as a leave-one-out analysis and, therefore, we have opted for a group analysis where both populations showed a high correlation (*R* = 0.95 and 0.96 for infants and adults respectively) with also a high concordance (*ρ_c_* = 0.95 and 0.96 for infants and adults respectively). Overall, the accuracy for the 50% holdout validation was below ± 4 mmHg for both populations. We note that, in the future, an external validation on a more heterogeneous cohort of patients should be used for the validation of the network to assess its real performance.

In comparison to our method, several TCD based methods have been used for the non-invasive estimation of ICP with an overall accuracy of ± 12 mmHg.^7^ Correlating the TCD pulsatility index with ICP, as shown in Bellner et al.,^42^ led to an accuracy of ± 4 mmHg, while other studies could not reproduce these results. Kim et al.^21^ used a semi-supervised method to analyze the morphology of the TCD waveform in order to classify intracranial hypertension, but this method was unable to give a continuous ICP estimate. Other emerging non-invasive methods either analyze acoustic signals which propagate through the cranium yielding in an accuracy of ± 6.8 mmHg to estimate ICP^43^ or measure with ultrasound or other imaging techniques the optic nerve sheath diameter to detect elevated ICP.^44^ Our results show a “best-case-scenario”, due to our use of all subjects in the training data-set, that has converged to a similar accuracy.

As a side note, Zacchetti et al.^45^ analyzed the accuracy of intraparenchymal ICP sensors in comparison to a ventricular catheter with an external transducer in a meta-analysis, concluding that using the random standard error the mean difference of 1.5 mmHg is acceptably small, but its accuracy of 11.4 mmHg shows a wide spread. This is an example illustrating the accuracy of invasive ICP sensors compared to each other, demonstrating once more the potential of our method.

As previously mentioned, there are other non-invasive ICP methods based on optics. In comparison to those, we were able to directly obtain an ICP value without the need of using surrogates like the CrCP^19^ or the need to indirectly look at metabolic alterations as was done with NIRS.^10,11^ In the recent preliminary results of Baker et al.,^19^ they have shown a good correlation of the estimated CrCP to ICP values in infants suffering from hydrocephalus. However, the CrCP was estimated before and after a shunt was placed and the ICP was measured at the moment the shunt was placed. Furthermore, continuous measurements of the CrCP are difficult since a continuous ABP signal is required. Contrary, our method does not rely on a second input like the ABP. Instead, it strongly depends on the learned features and the information which has provided the network for training. All in all, it shares with the other optical methods its advantage and suitability for continuous long term measurements at the bed site, while in other methods, e.g. using TCD, the head gear to hold the probes in place might be uncomfortable over time.

We admit that the accuracy of ± 4 mmHg in the random sample splitting method may be deceivingly good. This is because the training set partially contains information about all subjects used. Therefore, the overall bias is almost zero and negligible. Again, ideally, a larger data-set with more subjects and longer monitoring periods would enable a complete leave-one-out analysis for a true evaluation of the performance of the RNN.

The fact that neural networks can learn patterns from data without prior analysis via feature engineering makes it a strong algorithm in the engineering sense, especially, when it is not so clear what features from raw data may be most relevant. However, the concern with using neural networks on raw data in this sense is that if the problem turns out to be very complex, very large data-sets would be required for the algorithm to reach a point that it generalizes well. Besides training on a larger dataset, the extraction of specific features to reduce the dimensionality of the raw data could aid improving the model to generalize better for a dataset with a small sample size. Some pertinent features in pulsatile blood flow measured by DCS have recently been suggested by Ruesch et al.^46^ as valuable markers using a similar approach based on a random regression forest in a non-human primate model. On the other hand, there are other types of algorithms, such as convolutional neural networks (CNN)^47^ which could be implemented by integrating a CNN architecture to focus on local temporal correlations alongside an RNN analyzing complex feedback loops in the pulsatile CBF signal to prove a more fruitful method.

Ideally, such an algorithm should be trained on a large cohort covering the entire ICP range equally. Thus the method can be used directly as a stand-alone non-invasive technique to continuously predict an ICP index. However, if the limitations persist, one can think about the following possible clinical forecasting scenario. For example, all the gathered data is used to initially train the network. In a second stage, for each subject a short period of time is used to calibrate the method by training the pre-trained model with a measurement alongside an invasive monitor. With this procedure, clinicians can obtain information even after removing the invasive ICP sensor. One can also imagine calibrating the method with an invasive ICP measurement during surgery or by using a lumbar puncture. For this purpose, we have tried to test the algorithm using even less data for the training. By using randomly sampled 30% (≙ 70% holdout) and 10% (≙ 90% holdout) for the training we have achieved similar results compared to the 50% split analysis with a lowest accuracy of ± 4.7 mmHg. This shows clearly the potential for such a scenario, but further investigations especially for long term stability of the model are still needed.

Our data opens a window of opportunity to explore our algorithm in larger cohort of patients ideally enrolled in multi-center studies with better controlled conditions like using the same ICP sensors and protocols to gather enough data for generalization of the method.

## Conclusion

We have introduced and demonstrated a promising new method and an algorithm to estimate ICP in a continuous, non-invasive manner with DCS. In particular, a neural network was used to learn complex dynamics and features from pulsatile CBF measured optically by DCS to predict ICP values coming from an invasive ICP sensor. The method was demonstrated on infants with BESS and adults with TBI showing its versatility.

If the method is improved and validated on larger studies, it may have implications for both pediatrics as well as adults suffering from severe brain injury or hydrocephalus. It may additionally open new scenarios for patients with potential disruptions of ICP but not in a way to accept the risks of invasive ICP monitoring, such as those with ischemic strokes, unexplained headaches and others.

## Data Availability

Data available on request from the authors (JF, TD).

## Acknowledgments

We gratefully acknowledge valuable discussions with Wesley B. Baker, Daniel J. Licht and Arjun G. Yodh.

## Footnotes

a A 55 month old infant was enrolled as an *ad hoc* addition to the protocol as judged by the clinicians.

